# A Systematic Review of COVID - 19 Induced Myocarditis - Symptomatology, Prognosis, and Clinical Findings

**DOI:** 10.1101/2021.05.29.21258059

**Authors:** Vikash Jaiswal, Shavy Nagpal, Christine Angela E. Labitag, Janelle Tayo, Abhinav Patel, Kevin Bryan Lo, Rupalakshmi Vijayan, Wanessa F Matos, Sadia Yaqoob, Priyanka Panday, Saloni Savani, Zeinab Alnahas, Arushee Bhatnagar, Yoandra Diaz, John R. Dylewski

## Abstract

**Objective:** With the advent of a novel coronavirus in December 2019, several case studies have reported its adversity on cardiac cells. We conducted a systematic review that describes the symptomatology, prognosis, and clinical findings of patients with COVID-19-related myocarditis.

**Methods:** Search engines including PubMed, Google Scholar, Cochrane Central, and Web of Science were queried for “SARS-CoV-2” or “COVID 19” and “myocarditis.” PRISMA guidelines were employed, and peer-reviewed journals in English related to COVID-19 were included.

**Results:** This systematic review included 22 studies and 37 patients. Eight patients (36%) were confirmed myocarditis, while the rest were possible myocarditis. Most patients had elevated cardiac biomarkers, including troponin, CRP, CK, CK-MB, and NT-pro BNP. Electrocardiogram results noted tachycardia (47%), left ventricular hypertrophy (50%), ST-segment alterations (41%), and T wave inversion (18%). Echocardiography presented reduced LVEF (77%), left ventricle abnormalities (34%), right ventricle aberrations (12%), and pericardial effusion (71%). Further, CMR showed reduced myocardial edema (75%), non-ischemic patterns (50%), and hypokinesis (26%). The mortality was significant at 25%.

**Conclusions:** Mortality associated with COVID-19 myocarditis appears significant but underestimated. Further studies are warranted to evaluate and quantify patients’ actual prognosis and outcomes with COVID-19 myocarditis.

## 1 Introduction

On 30 April 2021, there have been 149,910,744 confirmed cases of COVID-19, including 3,155,168 deaths reported globally^1^. COVID-19 is known to target multiple organ systems, inclusive of the respiratory tract. The Angiotensin-converting enzyme 2 (ACE2) receptor has been hypothesized as the receptor used by the receptor-binding domain of virus surface spike protein to enter the host cell. Such affinity could be why the lungs are affected most compared to other organs, as there is a high concentration of ACE2 receptors on the lung epithelial surface^2^. On the other hand, pericytes lining the microvasculature have the strongest ACE2 receptor expression in the heart. Furthermore, the receptor is also expressed in cardiomyocytes, fibroblasts, and vascular smooth muscle cells, explaining why up to 28% of COVID-19 patients harbor myocardial injury^3,4^.

Myocarditis, the inflammation of the myocardial tissue, has been identified as the major cause of myocardial injury in COVID-19. Myocarditis is perceived as an increased immune response (cellular or humoral) in cardiac cells. Clinical manifestations range from chest pain, dyspnea to life-threatening conditions such as cardiogenic shock and death^5^. Myocarditis is confirmed by laboratory testing, ECG, cardiac imaging, heart catheterization, and endomyocardial biopsies (EMB)^6^. So far, several case studies have been reported. A meta–summary of cases presenting coronavirus-induced myocarditis identified chest pain, ECG changes, elevated cardiac and inflammatory biomarkers, left ventricular dysfunction, and hypokinesis as hallmark features of COVID-19-related myocarditis^7^. Using the findings from earlier reports, we have created a conceptual framework of the pathophysiology of COVID-19-related-myocarditis (Figure 1). It highlights the entry of the SARS-COV-2 virus to the heart and the signs and symptoms associated with the cardiac injury. Nevertheless, considering the limitations of published data (< 1 year of onset of COVID -19), more data is required to establish the symptomatology, prognosis, and clinical findings of COVID-19-related myocarditis. Thus, we further discussed COVID–19 induced myocarditis and provided an in-depth review of its symptomatology.

**Figure 1:**
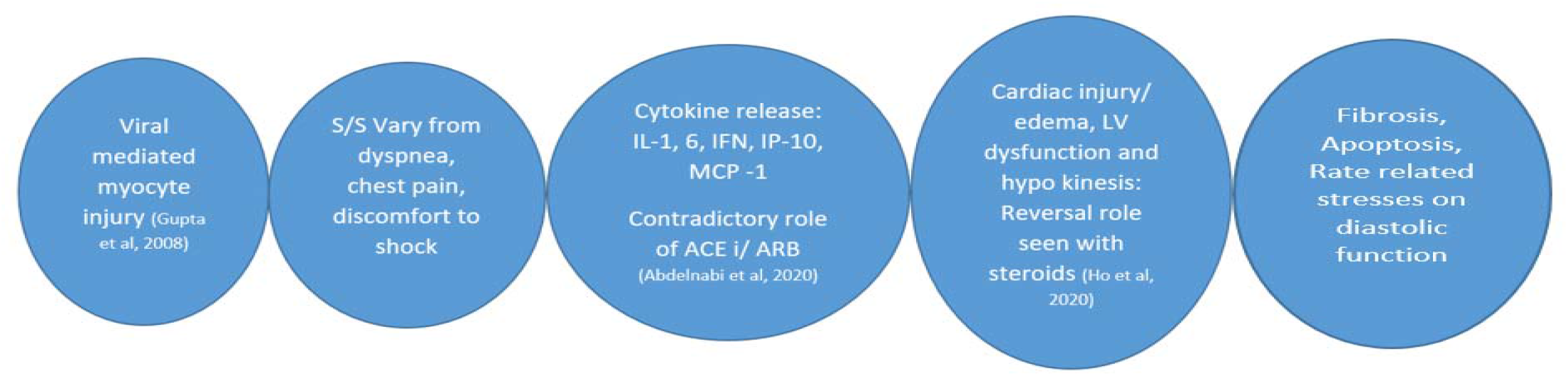
Snapshot: Myocarditis (COVID -19)

## 2 Methods

### 2.1 Protocol and Registration

We have used the PRISMA guidelines (Fig. 2) to identify eligible articles in this review.

**Figure 2:**
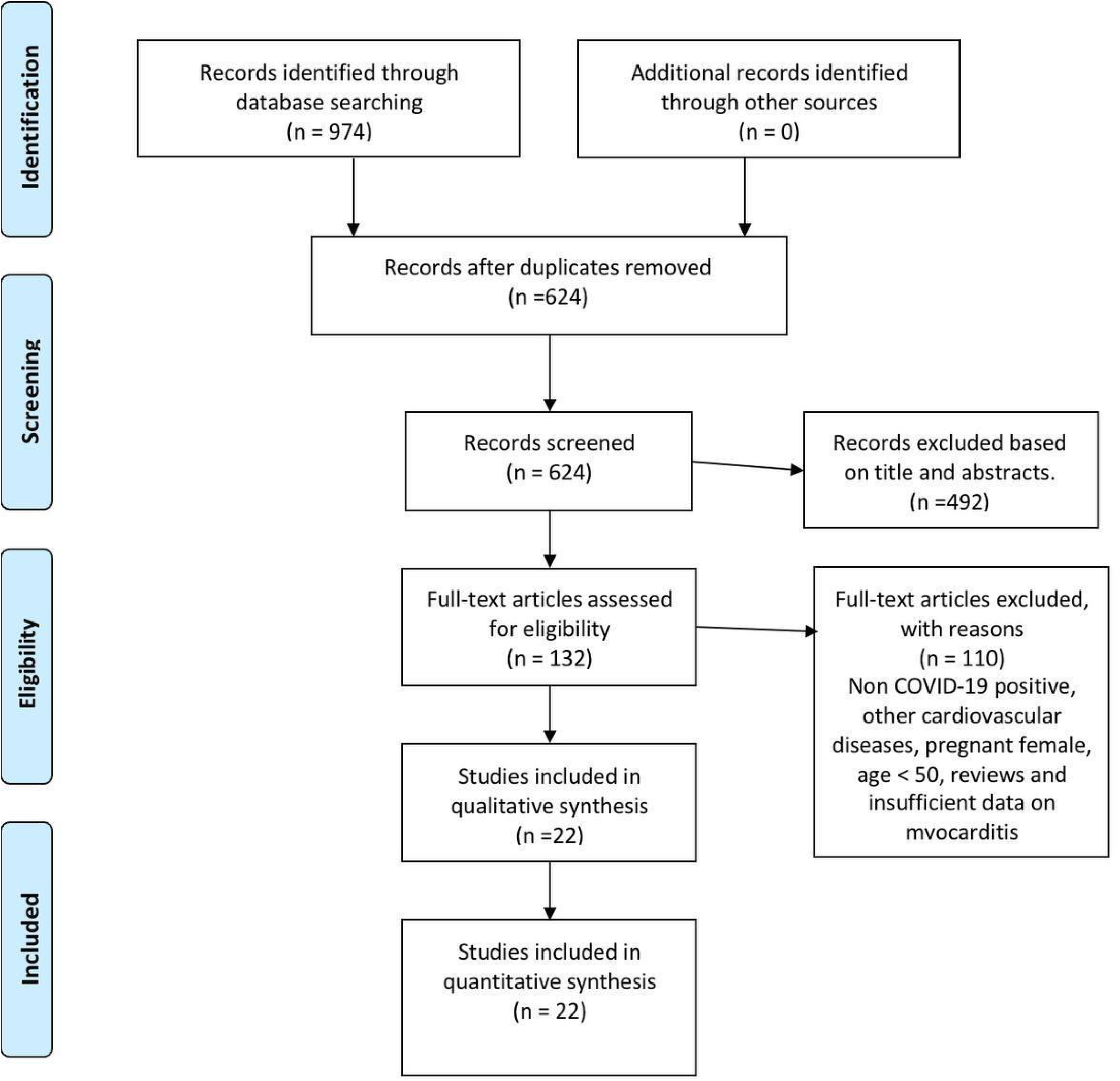
Flowchart on study selection and screening using PRISMA Guidelines.

### 2.2 Eligibility Criteria

#### A Inclusion Criteria

We included articles from peer-reviewed journals that have reported on COVID-19-induced myocarditis. It was limited to studies published only in English.

#### B Exclusion Criteria

Studies like systematic review, meta-analysis, and letter to the editor were excluded. We also excluded studies that include patients younger than 50 years old and those with known heart problems.

### 2.3 Information Sources and Search Strategies

A comprehensive literature search was done using the search engines Pubmed, Google Scholar, Cochrane CENTRAL, and Web of Science database. The search terms were “SARS-CoV-2” or “COVID 19” and “myocarditis.”

### 2.4 Study Selection

Two authors carried out an independent search and screened the titles and abstracts of the identified articles for inclusion. Afterward, full-text articles were reviewed to validate if they truly satisfy the inclusion criteria of this review. Any discrepancies were resolved by consulting a third author.

### 2.5 Data Collection Process and Data Items

Data extracted from articles included author/s, study design, number of patients, year of publication, country, setting, age in years, gender, comorbidities, symptoms, mortality, ECG findings, echocardiogram findings, laboratory results, imaging tests results, and myocardial biopsy. The data were analyzed and synthesized qualitatively using MS Excel PIVOT.

The quality of the included studies was rated using the ‘Research and Quality Scoring Method’ by Sackett and Haynes, the Jadad scale, and the items published by Cho and Bero^8^. Nine criteria were appraised to determine the overall quality of each study. The score ranged from zero to nine. Those with scores of zero to three were considered low quality, scores of four to six were moderate quality, and seven to nine were considered high quality. (Supplementary Table 1 and 2)

**Table 1:** Characteristics of the Included Studies

| Author | Study Design | Number of patients | Year of Publication | Country | Setting | Age in Years | Gender | Comorbidities |
| --- | --- | --- | --- | --- | --- | --- | --- | --- |
| Cizgici AY et al | Case report | 1 | 2020 | Turkey | ICU | 78 | Male | HTN |
| Yokoo P et al | Case report | 1 | 2020 | Brazil | Not specified | 81 | Male | HTN, ischemic stroke |
| Pietsch H et al | Case report | 1 | 2020 | Germany | ICU | 59 | Female | None |
| Pavon AG et al | Case report | 1 | 2020 | Switzerland | ICU | 64 | Male | Isolated pulmonary sarcoidosis and epilepsy |
| Khatrri A et al | Case report | 1 | 2020 | USA | ICU | 50 | Male | HTN, ischemic stroke |
| Hussain H et al | Case report | 1 | 2020 | USA | Isolation unit | 51 | Male | HTN |
| Dalen H et al | Case report | 1 | 2020 | Norway | ICU | 55 | Female | None |
| Zengh JH | Case report | 1 | 2020 | China | ICU | 63 | Male | None |
| Doyen D et al | Case report | 1 | 2020 | France | ICU | 69 | Male | HTN |
| Faircloth E et al | Case report | 1 | 2020 | USA | Not specified | 60 | Male | Multiple sclerosis |
| Coyle J | Case report | 1 | 2020 | USA | ED | 57 | Male | HTN |
| Luetkens JA et al, 2020 | Case report | 1 | 2020 | Germany | ICU | 79 | Male | Asthma |
| Jain A et al | Case report | 1 | 2020 | India | ED | 60 | Male | HTN, DM II |
| Mustafa S et al. | Case report | 1 | 2020 | USA | ED | 56 | Male | None |
| Mansoor A et al. | Case report | 1 | 2020 | USA | ED | 72 | Female | HTN |
| Al-assaf O et al. | Case report | 1 | 2020 | UAE | ED | 58 | Male | HTN |
| Khalid Y et al | Case report | 1 | 2020 | USA | Not specified | 76 | Female | HTN, hyperlipidemia, hypothyroidism |
| Ng My et al | Retrospective study | 16 | 2020 | China | ED | 53 to 69 | 7 Females and 9 Males | None |
| Inciardi et al | Case report | 1 | 2020 | Italy | ED | 53 | Female | None |
| Fried et al | Case report | 1 | 2020 | USA | Not specified | 64 | Female | HTN, hyperlipidemia |
| Wehit et al | Case report | 1 | 2020 | Argentina | ICU | 68 | Male | HTN, obesity, DM II, chronic smoking |
| Radbel et al | Case report | 1 | 2020 | USA | Not specified | 69 | female | DM II, RA, aplastic anemia |

Note: HTN – hypertension, DM II – type II diabetes mellitus, RA – rheumatoid *arthritis*

**Table number a2:** Signs and symptoms of COVID-19-related myocarditis

| Author | Systemic | Cardiovascular | Respiratory | Neurology | Gastrointestinal | Mortality |
| --- | --- | --- | --- | --- | --- | --- |
| Cizgici AY et al | N/A | Chest pain | Shortness of breath, ARDS | N/A | N/A | N/A |
| Yokoo P et al | Fever | N/A | Shortness of breath, No hypoxia (91% SpO2) | N/A | N/A | Recovered |
| Pietsch H et al | N/A | N/A | ARDS | N/A | N/A | N/A |
| Pavon AG et al | Fever (39.3) | Chest pain | Shortness of breath, Cough | N/A | N/A | Recovered |
| Khatiri A et al | Fever, Chills, Malaise | N/A | Shortness of Breath, Cough | Syncope | N/A | Yes |
| Hussain H et al | Fatigue, Fever | N/A | Cough, Shortness of breath | N/A | N/A | Recovered |
| Dalen H et al | Fatigue, Myalgia | Chest pain | Not mentioned | Syncope | N/A | Recovered |
| Zengh JH | Fever | N/A | Cough, Shortness of breath, Chest Pain | N/A | N/A | Yes |
| Doyen D et al | Fever | N/A | Cough, Shortness of breath | N/A | Vomiting, Diarrhea | Recovered |
| Faircloth E et al | Fever | Tachycardia, Hypotension | Shortness of breath, Tachypnea, Hypoxia | N/A | N/A | Recovered |
| Coyle J | Fever, Myalgia | N/A | Cough, Shortness of breath | N/A | Decrease appetite, Nausea, Diarrhea | Recovered |
| Luetkens JA et al, 2020 | Fatigue | N/A | Shortness of breath, No hypoxia (94% SpO2), Wheeze | Syncope | N/A | N/A |
| Jain A et al | N/A | N/A | Cough, Shortness of breath, hypoxia (75% SpO2) | N/A | N/A | Recovered |
| Mustafa S et al. | Fatigue, Myalgia | Chest pain | Cough, Shortness of breath | N/A | N/A | Recovered |
| Mansoor A et al. | Myalgia, Fever | Tachycardia | Cough, Colds, Tachypnea, Hypoxia (60% SpO2) | N/A | N/A | Yes |
| Al-assaf O et al. | N/A | Bradycardia | No hypoxia (98% SpO2) | N/A | N/A | Recovered |
| Khalid Y et al | Fever | Tachycardia | Cough, Shortness of breath, ARDS, Tachypnea, Hypoxia (79%SpO2) | N/A | N/A | N/A |
| Ng My et al | N/A | Chest pain | Cough, Shortness of breath, | N/A | N/A | N/A |
| Inciardi et al | Fatigue, Fever | Hypotension | Cough | N/A | N/A | Recovered |
| Fried et al | No fever | No chest pain | No shortness of breath | N/A | N/A | Recovered |
| Wehit et al | Fever, Fatigue | N/A | ARDS | N/A | N/A | N/A |
| Radbel et al | Fever, fatigue, Septic shock | Chest pain | Cough, ARDS | N/A | Abdominal pain | Yes |

## 3. Results

### 3.1. Study Selection

The electronic search identified 974 potential studies. No additional studies were obtained using other sources. Most of the articles were duplicates; hence, only 624 articles were screened initially. The title and abstracts were reviewed against the inclusion criteria, and 492 articles were excluded on primary screening. Around 110 articles were not on COVID-19-related myocarditis, did not provide an English translation, were review articles, pregnant female and involved participants <50 years old and with known heart problems were excluded. A review of the full-text manuscript of the 22 articles revealed that all of them met the eligibility criteria. Hence, all 22 articles^9-30^ were included in the systematic review illustrated according to PRISMA guidelines (Figure 2).

### 3.2. Characteristics of the Selected Studies

Twenty-two studies were selected for this systematic review, out of which twenty-one were case reports, and one was a retrospective study. All articles were published in the year 2020, and 41% were done within the US.

The reports included a total sample of 37 patients with COVID-19-related myocarditis. Cases were reported by patients aged between 50 to 81 years with male predominance (62%). The patients were primarily seen in the intensive care unit (41%) and emergency department (32%). In six studies (27%), the participants had no existing illness. In contrast, among those with existing morbidities, the most common diseases were hypertension (55%) and ischemic stroke (9%) (Table 1)

### 3.3 Methodological Quality of the Selected Studies

The ‘Research and Quality Scoring Method’ by Sackett and Haynes, the Jadad scale, and the items published by Cho and Bero were employed to appraise the quality of each study (Han et al., 2011)^8^. Out of 22 studies, 21 studies (96%) were deemed moderate, and one study (4%) has poor quality. The study design is a potential source of bias. The majority were case studies (96%), and there was one retrospective study (4%). Given the inherent limitations of case studies, the sample size was small, and there were no inclusion or exclusion criteria. Similarly, the sample size for the retrospective study was small at 16 (*S*upplementary Tables 1 and 2 presents the findings and the description of each criterion).

### 3.4 Results from the Selected Studies

#### Signs and Symptoms

Out of 22 studies, 17 articles reported the typical systemic symptoms of COVID-19 myocarditis, which included fever (77%), fatigue (41%), and myalgia (18%). Only 12 studies noted the cardiovascular complaints of the patients, such as chest pain (50%), tachycardia (25%), and hypotension (17%). Meanwhile, 21 studies discussed respiratory signs and symptoms, including shortness of breath (67%), cough (62%), acute respiratory distress syndrome (24%), hypoxia (19%), and tachypnea (14%). Out of 22 studies, only three reports examined the neurological presentation of COVID-19 myocarditis. All studies noted syncope (100%). Then, three studies documented the gastrointestinal complaints of the patients, wherein diarrhea (67%) was the most common presentation (Table 2).

#### Prognosis

Out of 22 studies, only 16 articles (73%) reported mortality, while six studies (27%) did not mention any consequent prognosis. Variable outcomes were reported on COVID-19-related myocarditis with a mortality of 25% (*n* = 4). Out of 16 patients, 12 of them (75%) recovered (Table 2).

#### Laboratory Finding

Most of the studies noted an increase in troponin (86%), NT-pro BNP (91%), ferritin (80%), WBC (80%), and D-dimer (67%) levels. The median troponin level was 290 (IQR = 3,543) ng/L, median NT-pro BNP was 4,639 (IQR = 4,678) pg/mL, median ferritin level was 948 (IQR = 244) ng/ml, mean WBC count was 17,500 (*SD* = 5,710.08) per μL, and median D-dimer level was 949 (IQR = 742) ng/mL. Meanwhile, in the studies that examined creatine, 50% noted risen levels with a mean creatinine level of 1.31 (*SD* = 0.48) mg/dL. On the other hand, all studies that evaluated CRP, CK level, CK-MB, ESR and procalcitonin noted elevated laboratory findings. The median CRP level was 18.01 (IQR = 14.09) mg/dL, median CK level was 1130 (IQR = 1005) U/L, median CK-MB level was 20.1 (IQR = 11.94) ng/ml, mean ESR was 80 (*SD* = 2.83) mm/hr, and median procalcitonin level was 7.69 (IQR = 7.51) ng/ml (Table 3)

**Table 3:**
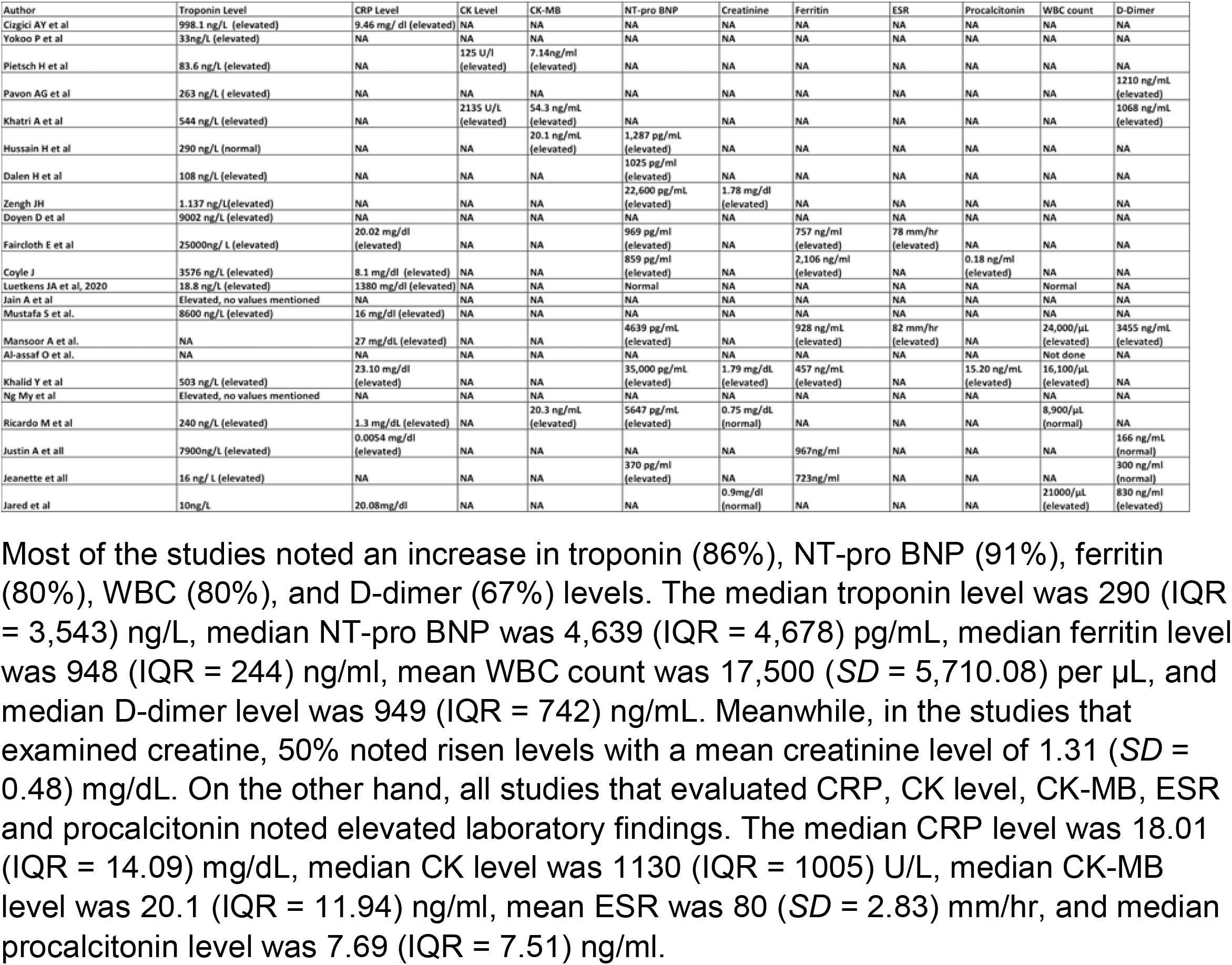
Laboratory findings of COVID-19-related myocarditis

#### Electrocardiogram

Tachycardia was noted in eight studies (47%). In terms of rhythm, normal sinus rhythm was noted in nine studies (75%) and atrial fibrillation in one study (8%). The ECG reports also revealed left ventricular hypertrophy (50%). The findings also noted alterations in the ST segments (41%) and T wave inversion (18%) (Table 4). All in all, these electrocardiogram findings vary pretty broadly.

**Table 4:** ECG findings of COVID-19 related myocarditis

| Author | Rate | Rhythm | Hypertrophy | Ischemia and Infarction | Miscellaneous |
| --- | --- | --- | --- | --- | --- |
| Cizgici AY et al | Tachycardia | fibrillation | NA | Concave ST elevation except for aVR lead | NA |
| Yokoo P et al | Normal | Normal | Normal | Normal | Normal |
| Pietsch H et al | NA | NA | NA | NA | NA |
| Pavon AG et al | NA | NA | NA | NA | NA |
| Khatri A et al | Tachycardia | Sinus Rhythm | NA | ST-elevation in leads II, III, aVF and ST-depression in I, aVL | NA |
| Hussain H et al | NA | NA | NA | Extensive and diffuse ST-segment elevation | NA |
| Dalen H et al | Tachycardia | Sinus Rhythm | NA | Insignificant ST-elevation in inferior leads, T-wave inversion in precordial leads. | Low-voltage ECG with peak-to-peak QRS amplitude less than 5 mm in the standard leads and 10 mm in the precordial leads (V5 and V6) |
| Zengh JH | Tachycardia | Sinus Rhythm | NA | No ST-segment elevation | NA |
| Doyen D et al | NA | NA | Left ventricular hypertrophy (LVH) | Diffuse inverted T waves in anterior leads | NA |
| Faircloth E et al | NA | NA | NA | Non-specific ST-changes in the anterior leads not consistent with STEMI | NA |
| Coyle J | Tachycardia | Sinus Rhythm | NA | No ST-T wave changes | NA |
| Luetkens JA et al, 2020 | Normal | Normal | Normal | Normal | Normal |
| Jain A et al | NA | NA | NA | Inferior infarct versus left anterior fascicular block; | Myocardial infarction findings were no longer present. |
| Mustafa S et al. | NA | NA | NA | ST elevations in the antero-lateral distribution | NA |
| Mansoor A et al. | Tachycardia | Sinus Rhythm | NA | Ischemic ST changes and fusion beats | PR elevation present in aVR and PR depression in leads II and aVF consistent with myopericarditis |
| Al-assaf O et al. | Bradycardia | Sinus Rhythm | NA | None | Repeat ECG 1 hour later showed 2:1 second-degree AV block |
| Khalid Y et al | Normal Rate | Sinus Rhythm | Left ventricular hypertrophy (LVH) | Normal | Short PR interval of 72 ms., and a QTc interval of 680 ms |
| Ng My et al | NA | NA | NA | NA | NA |
| Inciardi et al | Normal | Sinus Rhythm | NA | Minimal diffuse ST-segment elevation and an ST-segment depression with T-wave inversion in lead V1 and aVR | Low voltage in the limb leads |
| Fried et al | Tachycardia | Sinus Rhythm | NA | ST segment elevations in leads I, II, aVL, V2-V6, and PR elevation and ST depressions in aVR | Low voltage QRS complexes in the limb leads |
| Wehit et al | Normal | NA | NA | NA | NA |
| Radbel et al | Tachycardia | NA | NA | NA | NA |

#### Echocardiogram

The majority of studies (77%) observed a reduced left ventricular ejection fraction (LVEF). The mean LVEF was 33% (*SD* = 8.35). Out of the 17 studies, there were also significant abnormalities in the left ventricle in six studies (34%) and right ventricle in two studies (12%). Pericardial effusion was noted in 5 studies, but most studies had no mention of this (Table 5).

**Table 5:**
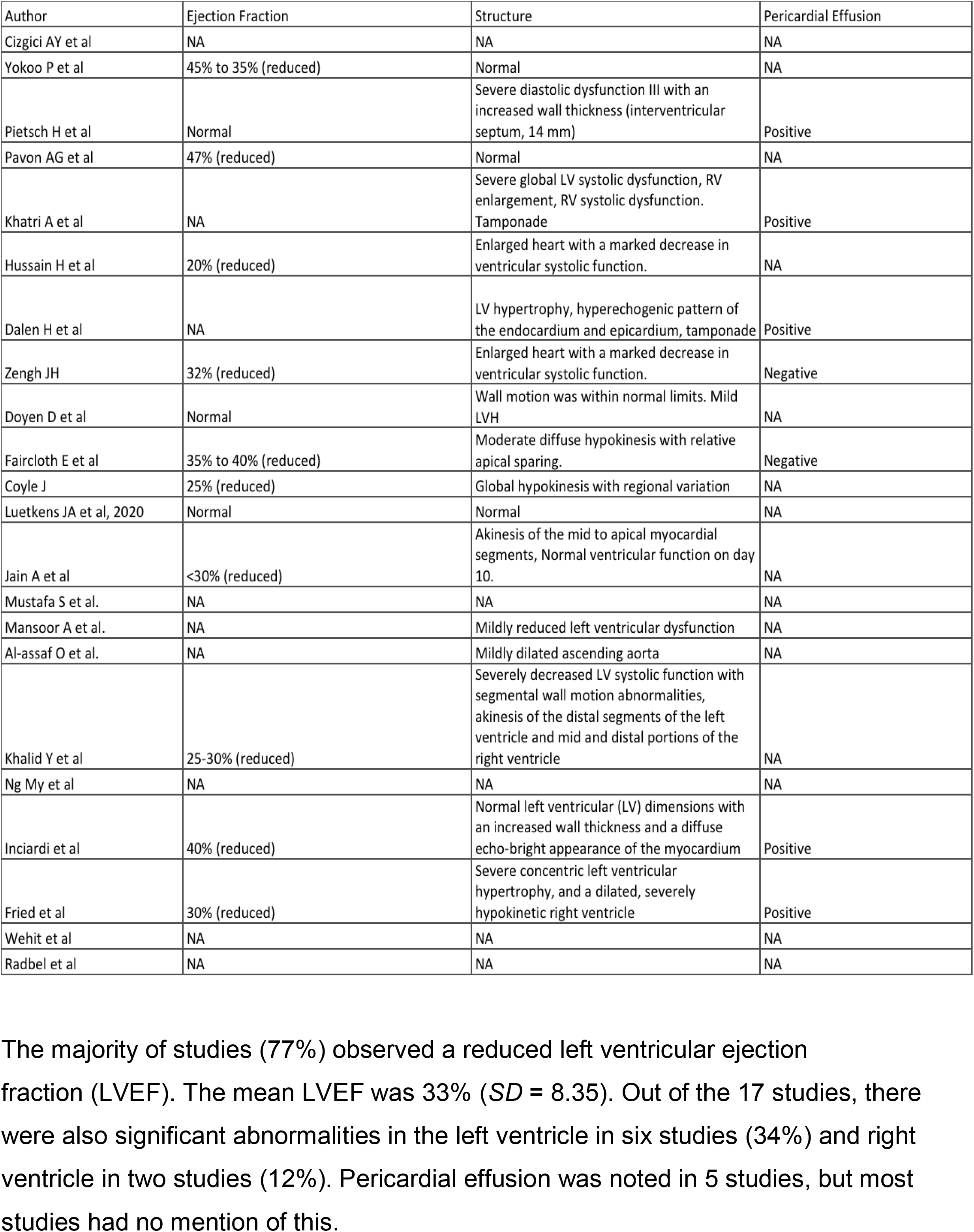
Echocardiogram findings of COVID-19 related myocarditis

#### Radiology

In terms of the coronary angiography results, only 1 study found a significant coronary artery disease, while of the ten studies, 8 (80%) had a chest CT noted with ground-glass changes, and 2 (20%) was noted with bilateral opacities. Similarly, in 14 studies with chest x-ray findings, 6 (43%) were noted with bilateral interstitial opacities, 2 (14%) with bilateral basal opacities, and 2 (14%) with bilateral pleural effusion (Table 6).

**Table 6:** Imaging findings of COVID-19 related myocarditis

| Author | Coronary Angiography | Chest CT | Chest x-ray |
| --- | --- | --- | --- |
| Cizgici AY et al | Negative for CAD | Ground glass changes | N/A |
| Yokoo P et al | N/A | Ground glass changes | N/A |
| Pietsch H et al | N/A | N/A | N/A |
| Pavon AG et al | N/A | N/A | N/A |
| Khatri A et al | Negative for CAD | Ground glass changes | N/A |
| Hussain H et al | Negative for CAD | Ground glass changes | Bilateral interstitial opacities |
| Dalen H et al | Negative for CAD | Ground glass changes | Bilateral pleural effusion |
| Zengh JH | N/A | N/A | Bilateral pleural effusion |
| Doyen D et al | N/A | Ground glass changes | Ground glass changes |
| Faircloth E et al | Negative CAD | Ground glass changes | N/A |
| Coyle J | N/A | N/A | Bilateral interstitial opacities |
| Luetkens JA et al, 2020 | N/A | Bilateral interstitial opacities | Bilateral interstitial opacities |
| Jain A et al | N/A | Ground glass changes | Normal |
| Mustafa S et al. | N/A | N/A | Bilateral interstitial opacities |
| Mansoor A et al. | N/A | N/A | Bilateral interstitial opacities |
| Al-assaf O et al. | Positive for CAD | N/A | Normal |
| Khalid Y et al | N/A | N/A | Bilateral interstitial opacities |
| Ng My et al | N/A | N/A | N/A |
| Inciardi et al | Negative for CAD | N/A | Normal |
| Fried et al | Negative for CAD | N/A | Normal |
| Wehit et al | N/A | N/A | Right basal opacities |
| Radbel et al | N/A | Bilateral nodular opacities | N/A |

#### CMR and EMB

The CMR and myocardial biopsy results were used to identify possible and confirmed cases of COVID-19-related myocarditis. Out of 22 studies, eight studies performed CMR, and only two were able to provide histopathological findings through EMB. Eight were confirmed cases (36%), and the rest were suspected cases (Table 7).

**Table 7:** CMR and myocardial biopsy findings

| Author | CMR | Myocardial biopsy | Possible or Confirmed Case |
| --- | --- | --- | --- |
| Cizgici AY et al | NA | NA | suspected |
| Yokoo P et al | Late enhancement areas with an ischemic pattern on the left ventricle base septum wall, with diffuse hypokinesis, and global systolic function | NA | Confirmed |
| Pietsch H et al | NA | EMB: Intramyocardial inflammation with absence of signs of necrosis. Increased no. of CD45R0+ T memory cells (96.15 cells/mm2), CD3+ cells (20.54 cells/mm2), CD11a+ cells (24.36 cells/mm2), CD11b+ cells (91.56 cells/mm2), and CD54+ cells (area fraction 1.91%), histology: hypertrophied myocytes (diameter 31 µm) | Confirmed |
| Pavon AG et al | Reduced left-ventricular (LV) systolic function ( 42%), mild hypokinesia of the lateral wall. T2-mapping sequences showed myocardial edema (segmental T2 = 55-57 ms) | NA | Confirmed |
| Khatiri A et al | NA | NA | suspected |
| Hussain H et al | NA | NA | suspected |
| Dalen H et al | Acute perimyocarditis, moderate enhancement in the anterolateral wall | NA | Confirmed |
| Zeng JH | NA | NA | suspected |
| Doyen D et al | Subepicardial late gadolinium enhancement of the apex and inferolateral wall | No significant findings | Confirmed |
| Faircloth E et al | NA | NA | suspected |
| Coyle J | NA | NA | suspected |
| Luetkens JA et al, 2020 | Normal LV size, mild systolic dysfunction (LV EF: 49%) with discrete hypokinesis, and pericardial effusion around the LV wall (+. 10 mm), diffuse interstitial myocardial edema with increased T2 signal, diffuse myocardial inflammation. | NA | Confirmed |
| Jain A et al | NA | NA | suspected |
| Mustafa S et al. | NA | NA | suspected |
| Mansoor A et al. | NA | NA | suspected |
| Al-assaf O et al. | Edema of the interventricular septum indicative of myocarditis | NA | Confirmed |
| Khalid Y et al | NA | NA | suspected |
| Ng My et al | Non-ischemic LGE with elevated global T2-mapping values (57-62 ms) | NA | suspected |
| Inciardi et al | Increased wall thickness with diffuse biventricular apical hypokinesis, severe LV dysfunction (LVEF of 35%) , Short tau inversion recovery and T2-mapping sequences showed marked biventricular myocardial interstitial edema. | NA | Confirmed |
| Fried et al | NA | NA | suspected |
| Wehit et al | NA | NA | suspected |
| Radbel et al | NA | NA | suspected |

Generally, myocarditis can be suspected with clinical presentations suggestive of acute coronary syndrome on ECG, laboratory testing (e.g., increase troponin levels), and/or wall motion abnormalities with no obstruction of coronary arteries on coronary angiography^4^.

Myocarditis diagnosed through CMR was based on Lake Louise Criteria, which includes signs of edema, T2-weighted imaging, and necrosis with late gadolinium enhancement (LGE), along with supportive findings of left ventricular dysfunction and pericardial effusion^11^. On T1-weighted imaging, two studies revealed diffuse biventricular hypokinesis, one discrete hypokinesis, and one mild hypokinesis in the lateral wall segment. On T2 weighted imaging, six studies with T2 mapping of myocardium illustrated myocardial edema, of which 2 of them revealed a diffuse, biventricular and interventricular region. Imaging on LGE results showed non-ischemic patterns in 4 studies and an ischemic pattern on the left ventricular septum wall in 1 study. Anterolateral, inferolateral, and biventricular walls were the most affected regions with subepicardial to transmural affectation. Supporting criteria in the Lake Louise consensus criteria, such as systolic dysfunction, was also noted in 3 studies, with a mean value LVEF of 42% and pericardial effusion in 1 study.

From the two studies that performed EMB, 1 study showed an increased number of T-lymphocytes infiltrates^11^. The result of the other study excluded other potential etiologies such as heart failure or arrhythmias, parvovirus B19, human herpesvirus, Epstein-Barr virus, enterovirus, cytomegalovirus, adenovirus, HIV, and hepatitis C virus. They have concluded that SARS-CoV-2 infection was most likely the cause^17^.

## Discussion

This systematic review aimed to describe the symptomatology, prognosis, and clinical findings of patients with probable and confirmed COVID-19-related myocarditis. Frequent clinical findings of COVID-19 constitute fever, cough, shortness of breath, and fatigue^31^. WHO (World Health Organization) has also cited fever and cough as striking features of COVID-19^32^. Fever, dyspnea, and/or chest pain are the typical manifestations of myocarditis that overlap with the symptoms of COVID-19, thus making diagnosis challenging^33^. However, laboratory investigations such as raised cardiac biomarkers and electrocardiogram may assist in diagnosing COVID-19 induced myocarditis.

Our study’s mortality rate from COVID-19-related myocarditis was high as 25%, which is comparable with the previous meta-summary, which noted 27% fatalities in patients with COVID-19 related myocarditis^34^. Our results align with a prior case report which proposes that myocardial injury is a cardinal predictor of mortality in COVID-19^35^. Even though the majority of the patients in our review survived COVID-19-related myocarditis, the actual mortality rate may be higher as many of the included studies did not report a fatality.

COVID-19 symptoms can be minimal to severe^36^. This was evident in our sample, wherein the participants displayed a varied course of the disease. The immune system plays a vital role in the severity of the symptoms in an individual inflicted with COVID-19. Those individuals with strong immunity are found to have mild symptoms compared to those with weak immunity. In contrast, aged people who have multiple comorbidities show more signs and symptoms of COVID-19^37^. Some of these patients can progress to acute respiratory distress syndrome, a severe complication of COVID-19^38^.

Conventionally, serum biomarkers are used to confirm any suspected case of acute myocarditis. Patients infected with the COVID-19 exhibited elevated troponin levels^39^. However, their presence cannot be used to rule out the possibility of any specific types of myocarditis. High CK-MB and BNP levels may be associated with COVID-19 induced myocarditis but are relatively non-specific as any state of volume overload or demand ischemia may also present likewise^40^. However, inflammatory markers such as C-reactive protein, D-dimer, and ferritin are found to be significantly increased in severe illness^41^. ECG findings of included studies denoted tachycardia and left ventricular hypertrophy, which coincides with the study’s conceptual framework.

Furthermore, the echocardiogram findings were positive for a reduced left ventricular ejection fraction (LVEF) varying between 20% to 47% and pericardial effusion. Pericardial effusion appears to be a novel finding. Nevertheless, past literature on clinically suspected myocarditis and acute myocarditis recognized pericardial effusion as a concurrent finding in patients with myocarditis^42,43^. Nevertheless, reduced ejection fraction is not distinct in myocarditis. It can also occur in a myocardial injury secondary to systemic causes rather than a viral infection of the heart. Definitive findings can be ascertained in the endomyocardial biopsy and MRI, but we had a very small proportion of this cohort study. Despite such measures, we noticed mortality in one-fourth of cases during recovery in three-fourths. The authors addressed this as a limitation for this study.

## Limitations

COVID-19 and myocarditis can potentially present the same way and maybe hard to differentiate clinically. It may be underreported as many patients account for “new” diminished EF with elevated myocardial markers, but it is plausible that projected growth in severe outcomes might warrant special attention shortly. However, only a handful of myocarditis cases confirmed via MRI and/ or endomyocardial biopsy have been addressed. Patients were reluctant to undergo MRI and biopsy due to fear of contracting COVID-19, and available diagnostic tests such as echo and EKG were reliable for screening but not diagnostic purposes, except in the case of pericardial effusion. COVID-19 is associated with cardiovascular involvement, and patients with known cases of cardiovascular disease are prone to face severe outcomes with COVID-19. It is more toilsome to differentiate COVID-19 patients with and concomitant myocarditis based on clinical features. Biomarkers such as troponin, BNP, and CK-MB might aid in the diagnosis but are non-specific because their levels can also rise in other conditions such as acute heart failure and demand ischemia.

## Conclusions

COVID-19 related mortality from myocarditis appears significant and under-estimated. Many cases of COVID-19 myocarditis have not been subjected to definitive diagnostic approaches, including endomyocardial biopsy and MRI. In addition, rates of poor outcomes such as mortality and the presence of myocarditis itself might be underreported. Further studies are needed to outline the trends of symptoms, outcomes, and prognosis of patients with COVID-19 related myocarditis. Moreover, an upsurge in such cases necessitates establishing treatment guidelines since current treatment approaches in practice vary from case to case.

## Data Availability

available on request

## Abbreviations

S/S: signs and symptoms
HTN: hypertension
DM II: type II diabetes mellitus
RA: rheumatoid *arthritis*
NA-Not: available

## Author’s Role

1. **Research Project: A. Conception, B. Organization, C. Execution**.
2. **Statistical Analysis: A. Design, B. Execution, C. Review, and Critique**.
3. **Manuscript: A. Writing of the first draft, B. Review of Critique**.

**VJ: 1A, 1B, 1C, 2A, 2B, 2C, 3A, 3B**

**SN: 1B, 1C, 2C, 3A, 3B**

**JT, AP, CE: 1B, 1C, 2B, 2A, 3A**

**WM, ZA, YD, RV, SS, AB, PP: 2B, 2C, 3B**

**KB, SY: 1C, 2C, 3A, 3B**.

**JD: 2C, 3B**.

## Acknowledgment

The authors of this article would like to thanks the sponsoring institution “Larkin Community Hospital” for helping and supporting throughout till final submission. w

## References

1. World Health Organization (2021a). WHO Coronavirus disease (COVID-19) dashboard. https://covid19.who.int/.

2. Rothan, H. A., & Byrareddy, S. N. (2020). The epidemiology and pathogenesis of coronavirus disease (COVID-19) outbreak. Journal of Autoimmunity, 109 (102433). https://doi.org/10.1016/j.jaut.2020.102433.

3. Guo, T., Fan, Y., Chen, M., Wu, X., Zhang, L., He, T., Wang, H., Wan, J., Wang, X., & Lu, Z. (2020). Cardiovascular implications of fatal outcomes of patients with coronavirus Disease 2019 (COVID-19). JAMA Cardiology, 5(7), 811–818. https://doi.org/10.1001/jamacardio.2020.1017.

4. Tucker, N. R., Chaffin, M., Bedi Jr., K.C., Papangeli, I., Akkad, A.D., Arduini, A., Hayat, S., Eraslan, G., Muus, C., Bhattacharyya, R., Stegmann, C., Human Cell Atlas Lung Biological Network, Margulies, K.B., & Ellinor, P. (2020). Myocyte-specific upregulation of ACE2 in cardiovascular disease: Implications for SARS-COV-2–mediated myocarditis. Circulation, 142(7), 708–710. https://doi.org/10.1161/CIRCULATIONAHA.120.047911.

5. Tschöpe, C., Cooper, L., Torre-Amione, G., & Linthout, S.V. (2019). Management of myocarditis-related cardiomyopathy in adults. Circulation Research, 124(11), 1568–1583. https://doi.org/10.1161/CIRCRESAHA.118.313578.

6. Ukena, C., Kindermann, M., Mahfoud, F., Geisel, J., Lepper, P.M., Kandolf, R., Bohm, M., Kindermann, I. (2014). Diagnostic and prognostic validity of different biomarkers in patients with suspected myocarditis. Clinical Research in Cardiology. https://www.escardio.org/Working-groups/Working-Group-on-Myocardial-and-Pericardial-Diseases/Publications/Paper-of-the-Month/Diagnostic-and-prognostic-validity-of-different-biomarkers-in-patients-with-susp

7. Ho, J.S., Sia, C.H., Chan, M.Y., Lin, W., & Wong, R.C. (2020). Coronavirus-induced myocarditis: A meta-summary of cases. The Journal of Cardiopulmonary and Acute Care, 49(6), 681–685 https://doi.org/10.1016/j.hrtlng.2020.08.013.

8. Han, H.R., Kim, J., Lee, J.E., Hedlin, H.K., Song, H., Song, Y., & Kim, M.T. (2011). Interventions that increase use of Pap tests among ethnic minority women: A meta-analysis. Psycho-Oncology, 20, 341–351. https://doi.org/10.1002/pon.1754.

9. Cizgici, A. Y., Zencirkiran Agus, H., & Yildiz, M. (2020). COVID-19 myopericarditis: It should be kept in mind in today’s conditions. The American Journal of Emergency Medicine, 38 (7), 1547.e5–1547.e6. https://doi.org/10.1016/j.ajem.2020.04.080. Result section starts

10. Yokoo, P., Fonseca, E., Sasdelli Neto, R., Ishikawa, W. Y., Silva, M., Yanata, E., Chate, R. C., Nunes Filho, A., Bettega, M., Fernandes, J., Tarasoutchi, F., & Szarf, G. (2020). COVID-19 myocarditis: a case report. Einstein (Sao Paulo, Brazil), 18, eRC5876. https://doi.org/10.31744/einstein_journal/2020RC5876

11. Pietsch, H., Escher, F., Aleshcheva, G., Baumeier, C., Morawietz, L., Elsaesser, A., & Schultheiss, H. P. (2021). Proof of SARS-CoV-2 genomes in endomyocardial biopsy with latency after acute infection. International Journal of Infectious Diseases: Official Publication of The International Society for Infectious Diseases, 102, 70–72. https://doi.org/10.1016/j.ijid.2020.10.012.

12. Pavon, A. G., Meier, D., Samim, D., Rotzinger, D. C., Fournier, S., Marquis, P., Monney, P., Muller, O., & Schwitter, J. (2020). First documentation of persistent SARS-Cov-2 infection presenting with late acute severe myocarditis. The Canadian Journal of Cardiology, 36(8), 1326.e5–1326.e7. https://doi.org/10.1016/j.cjca.2020.06.005.

13. Khatri, A., & Wallach, F. (2020). Coronavirus disease 2019 (COVID-19) presenting as purulent fulminant myopericarditis and cardiac tamponade: A case report and literature review. Hear. & lung: The Journal of Critical Care, 49 (6), 858–863. https://doi.org/10.1016/j.hrtlng.2020.06.003.

14. Hussain, H., Fadel, A., Alwaeli, H., & Guardiola, V. (2020). Coronavirus (COVID-19) Fulminant myopericarditis and acute respiratory distress syndrome (ards) in a middle-aged male patient. Cureus, 12 (6), e8808. https://doi.org/10.7759/cureus.8808.

15. Dalen, H., Holte, E., Guldal, A. U., Hegvik, J. A., Stensaeth, K. H., Braaten, A. T., Mjølstad, O. C., Rossvoll, O., & Wiseth, R. (2020). Acute perimyocarditis with cardiac tamponade in COVID-19 infection without respiratory disease. BMJ Case Reports, 13 (8), e236218. https://doi.org/10.1136/bcr-2020-236218.

16. Zeng, J. H., Liu, Y. X., Yuan, J., Wang, F. X., Wu, W. B., Li, J. X., Wang, L. F., Gao, H., Wang, Y., Dong, C. F., Li, Y. J., Xie, X. J., Feng, C., & Liu, L. (2020). First case of COVID-19 complicated with fulminant myocarditis: A case report and insights. Infection, 48(5), 773–777. https://doi.org/10.1007/s15010-020-01424-5.

17. Doyen, D., Moceri, P., Ducreux, D., & Dellamonica, J. (2020). Myocarditis in a patient with COVID-19: A cause of raised troponin and ECG changes. Lancet (London, England), 395(10235), 1516. https://doi.org/10.1016/S0140-6736(20)30912-0.

18. Faircloth, E., Conner, C., Dougherty, K., Arora, S., & Thorevska, N. (2020). Viral heartbreak: A case of covid-19 myocarditis vs stress-induced cardiomyopathy. Chest, 158 (4), A173. https://doi.org/10.1016/j.chest.2020.08.185.

19. Coyle, J., Igbinomwanhia, E., Sanchez-Nadales, A., Danciu, S., Chu, C., & Shah, N. (2020). A recovered case of COVID-19 myocarditis and ARDS treated with corticosteroids, tocilizumab, and experimental AT-001. JACC Case reports, 2(9), 1331–1336. https://doi.org/10.1016/j.jaccas.2020.04.025.

20. Luetkens, J. A., Isaak, A., Zimmer, S., Nattermann, J., Sprinkart, A. M., Boesecke, C., Rieke, G. J., Zachoval, C., Heine, A., Velten, M., & Duerr, G. D. (2020). Diffuse myocardial inflammation in COVID-19 associated myocarditis detected by multiparametric cardiac magnetic resonance imaging. Circulation. Cardiovascular imaging, 13 (5), e010897. https://doi.org/10.1161/CIRCIMAGING.120.010897.

21. Jain, A., Deval, N., & Paul, L. (2020). A recovered case of covid-19 myocarditis treated with IV immunoglobulin. Chest, 158 (4), A281. https://doi.org/10.1016/j.chest.2020.08.282.

22. Mustafa, S., Zafar, M., Agrawal, N., Shahbaz, A., & Al-khafaji, N. (2020). COVID-19-associated myocarditis mimicking ST elevation myocardial infarction. Chest, 158 (4), A572. https://doi.org/10.1016/j.chest.2020.08.537.

23. Mansoor, A., Chang, D., & Mitra, R. (2020). Rhythm, conduction, and ST elevation with COVID-19: Myocarditis or myocardial infarction? HeartRhythm Case Reports, 6 (10), 671–675. https://doi.org/10.1016/j.hrcr.2020.08.001.

24. Al-assaf, O., Mirza, M., & Musa, A. (2020). Atypical presentation of COVID-19 as subclinical myocarditis with persistent high-degree atrioventricular block treated with pacemaker implant. HeartRhythm Case Reports, 6 (11) 884–887. https://doi.org/10.1016/j.hrcr.2020.09.003.

25. Khalid, Y., Dasu, N., & Dasu, K. (2020). A case of novel coronavirus (COV8ID-19)-induced viral myocarditis mimicking a Takotsubo cardiomyopathy. HeartRhythm Case Reports, 6 (8), 473–476. https://doi.org/10.1016/j.hrcr.2020.05.020.

26. Ng, M. Y., Ferreira, V. M., Leung, S. T., Lee, J. C. Y., Fong, A. H. T., Liu, R. W. T, Chan, J. W. M., Ka Lun Wu, A., Lung, K. C., Crean, A., Fan-Ngai Hung, I., & Siu, C. W. (2020). Patients recovered from COVID-19 show ongoing subclinical myocarditis as revealed by cardiac magnetic resonance imaging. JACC: Cardiovascular Imaging, 13 (11), 2476–2478. https://doi.org/10.1016/j.jcmg.2020.08.012.

27. Inciardi, R. M., Lupi, L., Zaccone, G., Italia, L., Raffo, M., Tomasoni, D., Cani, D., Cerini, Manuel., Farina, Davide., Gavazzi, Emanuele., Maroldi, R., Adamo, M., Ammirati, E., Sinagra, G., Lombardi, C. M., & Metra, M. (2020). Cardiac involvement in a patient with coronavirus Disease 2019 (COVID-19). JAMA Cardiology, 5 (7), 819–824. https://doi:10.1001/jamacardio.2020.1096.

28. Fried, J. A., Ramasubbu, K., Bhatt, R., Topkara, V. K., Clerkin, K. J., Horn, E., Rabbani, L., Brodie, D., Jain, S. S., Kirtane, A. J., Masoumi, A., Takeda, K., Kumaraiah, D., Burkhoff, D., Leon, M., Schwartz, A., Uriel, N., & Sayer, G. (2020). The variety of cardiovascular presentations of COVID-19. Circulation, 141 (23), 1930–1936. https://doi.org/10.1161/CIRCULATIONAHA.120.047164.

29. Wehit, J., Sosa, F., Merlo, P., Roberti, J., Osatnik, J., (2020). Identification of COVID-19-associated myocarditis by speckle-tracking transesophageal echocardiography in critical care. Acta Colombiana de Cuidado Intensivo. https://doi.org/10.1016/j.acci.2020.11.008.

30. Radbel, J., Narayanan, N., & Bhatt, P. J. (2020). Use of tocilizumab for COVID-19-induced cytokine release syndrome: A cautionary case report. Chest, 158 (1), e15–e19. https://doi.org/10.1016/j.chest.2020.04.024

31. Fu L, Wang B, Yuan T, Chen X, Ao Y, Fitzpatrick T, Li P, Zhou Y, Lin Y, Duan Q, Luo G, Fan S, Lu Y, Feng A, Zhan Y, Liang B, Cai W, Zhang L, Du X, Li L, Shu Y, Zou H. Clinical characteristics of coronavirus disease 2019 (COVID-19) in China: a systematic review and meta-analysis. J Infect 2020; S0163-4453(20)30170-5.

32. Advice for the public on covid-19 – world health organization. Accessed May 6, 2021. https://www.who.int/emergencies/diseases/novel-coronavirus-2019/advice-for-public

33. Cooper LT Jr. Myocarditis. N Engl J Med 2009; 360(15): 1526–38.

34. Ho, J.S., Sia, C.H., Chan, M.Y., Lin, W., & Wong, R.C. (2020). Coronavirus-induced myocarditis: A meta-summary of cases. The Journal of Cardiopulmonary and Acute Care, 49(6), 681–685 https://doi.org/10.1016/j.hrtlng.2020.08.013.

35. Zhou, F., Yu, T., Du, R., Fan, G., Liu, Y., Liu, Z., Xiang, J.,Wang, Y., Song, B., Gu, X., Guan, L., Wei, Y., Li, H., Wu, X., Xu, J., Tu, S., Zhang, S., Chen, H., & Cao, B. (2020). Clinical course and risk factors for mortality of adult in patients with COVID-19 in Wuhan, China: A retrospective cohort study. Lancet, 395(10229), 1054–1062. https://doi.org/10.1016/S0140-6736 (20)30566-3.

36. Yuki, K., Fujiogi, M., & Koutsogiannaki, S. (2020). COVID-19 pathophysiology: A review. Clinical Immunology, 215, 108427. https://doi.org/10.1016/j.clim.2020.108427.

37. Brodin P. Immune determinants of COVID-19 disease presentation and severity. Nature Medicine. 2021 Jan;27(1):28–33. DOI: 10.1038/s41591-020-01202-8.

38. Hussain, H., Fadel, A., Alwaeli, H., & Guardiola, V. (2020). Coronavirus (COVID-19) Fulminant myopericarditis and acute respiratory distress syndrome (ards) in a middle-aged male patient. Cureus, 12 (6), e8808. https://doi.org/10.7759/cureus.8808.

39. Qin JJ, Cheng X, Zhou F, Lei F, Akolkar G, Cai J, Zhang XJ, Blet A, Xie J, Zhang P, Liu YM, Huang Z, Zhao LP, Lin L, Xia M, Chen MM, Song X, Bai L, Chen Z, Zhang X, Xiang D, Chen J, Xu Q, Ma X, Touyz RM, Gao C, Wang H, Liu L, Mao W, Luo P, Yan Y, Ye P, Chen M, Chen G, Zhu L, She ZG, Huang X, Yuan Y, Zhang BH, Wang Y, Liu PP, Li H. Redefining Cardiac Biomarkers in Predicting Mortality of Inpatients With COVID-19. Hypertension. 2020 Oct;76(4):1104–1112. doi: 10.1161/HYPERTENSIONAHA.120.15528. Epub 2020 Jul 14. PMID: 32673499; PMCID: PMC7375179.

40. Gaggin H.K. & Januzzi J.L. (2015). Cardiac Biomarkers and Heart Failure, Expert Analysis. American College of Cardiology. Feb,10, 2015.

41. Ho, J.S., Sia, C.H., Chan, M.Y., Lin, W., & Wong, R.C. (2020). Coronavirus-induced myocarditis: A meta-summary of cases. The Journal of Cardiopulmonary and Acute Care, 49(6), 681–685 https://doi.org/10.1016/j.hrtlng.2020.08.013.

42. Ukena, C., Kindermann, M., Mahfoud, F., Geisel, J., Lepper, P.M., Kandolf, R., Bohm, M., Kindermann, I. (2014). Diagnostic and prognostic validity of different biomarkers in patients with suspected myocarditis. Clinical Research in Cardiology. https://www.escardio.org/Working-groups/Working-Group-on-Myocardial-and-Pericardial-Diseases/Publications/Paper-of-the-Month/Diagnostic-and-prognostic-validity-of-different-biomarkers-in-patients-with-susp.

43. Di Bella, G., Imazio, M., Bogaert, J., Pizzino, F., Camastra, G., Monti, L., Dellegrottaglie, S., Donato, R., Moro, C., Pepe, A., Lanzillo, C., Pontone, G., Marra, M.P., Fusco, A., Scatteia, A., Pingitore, A., & Aquaro, G.D. (2019). Clinical value and prognostic impact of pericardial involvement in acute myocarditis. Circulation: Cardiovascular Imaging, 12(2). https://doi.org/10.1161/CIRCIMAGING.118.008504

