## Supplementary Table 1 for "A Systematic Review of COVID - 19 Induced Myocarditis - Symptomatology, Prognosis, and Clinical Findings"

***Supplementary Table 1 (Quality Appraisal)***

| **CRITERIA** | | Cizgici AY et al | | | Yokoo P et al | | Pietsch H et al | | Pavon AG et al | Khatri A et al | Hussain H et al | | Dalen H et al | Zengh JH |
| --- | --- | --- | --- | --- | --- | --- | --- | --- | --- | --- | --- | --- | --- | --- |
| Study design | | 0 | | | 0 | | 0 | | 0 | 0 | 0 | | 0 | 0 |
| Outcome measure | | 1 | | | 1 | | 1 | | 1 | 1 | 1 | | 1 | 1 |
| Clarity of outcome | | 1 | | | 1 | | 1 | | 1 | 1 | 1 | | 1 | 1 |
| Information on withdrawal or dropout rate | | 0 | | | 0 | | 0 | | 0 | 0 | 0 | | 0 | 0 |
| Research question | | 1 | | | 1 | | 1 | | 1 | 1 | 1 | | 1 | 1 |
| Participants in sample | | 1 | | | 1 | | 1 | | 1 | 1 | 1 | | 1 | 1 |
| Participant inclusion/ exclusion criteria | | 0 | | | 0 | | 0 | | 0 | 0 | 0 | | 0 | 0 |
| Type of location where study conducted and type of data collection performed | | 1 | | | 1 | | 1 | | 1 | 1 | 1 | | 1 | 1 |
| Sample size justification and power analysis | | 0 | | | 0 | | 0 | | 0 | 0 | 0 | | 0 | 0 |
| **TOTAL** | | 5 | | | 5 | | 5 | | 5 | 5 | 5 | | 5 | 5 |
| **RATING** | | Moderate | | | Moderate | | Moderate | | Moderate | Moderate | Moderate | | Moderate | |
| Doyen D et al | Faircloth E et al | | Coyle J | Luetkens JA et al | | Jain A et al | | Mustafa S et al. | Faircloth E et al. | Mansoor A et al. | Al-assaf O et al. | Khalid Y et al | Ng My et al | |
| 0 | 0 | | 0 | 0 | | 0 | | 0 | 0 | 0 | 0 | 0 | 0 | |
| 1 | 1 | | 1 | 1 | | 1 | | 1 | 1 | 1 | 1 | 1 | 1 | |
| 1 | 1 | | 1 | 1 | | 1 | | 1 | 1 | 1 | 1 | 1 | 0 | |
| 0 | 0 | | 0 | 0 | | 0 | | 0 | 0 | 0 | 0 | 0 | 0 | |
| 0 | 0 | | 1 | 1 | | 1 | | 1 | 0 | 1 | 1 | 1 | 1 | |
| 1 | 1 | | 1 | 1 | | 1 | | 1 | 1 | 1 | 1 | 0 | 1 | |
| 0 | 0 | | 0 | 0 | | 0 | | 0 | 0 | 0 | 0 | 0 | 1 | |
| 1 | 0 | | 1 | 1 | | 1 | | 1 | 0 | 1 | 1 | 1 | 1 | |
| 0 | 0 | | 0 | 0 | | 0 | | 0 | 0 | 0 | 0 | 0 | 0 | |
| 4 | 3 | | 5 | 5 | | 5 | | 5 | 3 | 5 | 5 | 4 | 5 | |
| Moderate | Low | | Moderate | Moderate | | Moderate | | Moderate | Low | Moderate | Moderate | Moderate | Moderate | |

**Supplementary table 2 (Scoring)**

| **CRITERIA** | **SCORING** |
| --- | --- |
| Study design | 0 = Non-randomized, systematic review, observational, cohort study |
|  | 1 = Randomized experiment, quasi-experiment |
| Outcome measure | 0 = Measure of the outcome is self report |
|  | 1 = Measure of the outcome mentioned using validated measure |
| Clarity of outcome | 0 = No definition of study outcome |
|  | 1 = Clearly defined the study outcome |
| Information on withdrawal or dropout rate | 0 = Not stated or discussed |
|  | 1 = Clearly stated [the number and the reasons for withdrawals in each group are stated] |
| Research question | 0 = Not clear |
|  | 1 = Clearly stated |
| Participants in sample | 0 = Not clear |
|  | 1 = Clearly identified and described |
| Participant inclusion/ exclusion criteria | 0 = Not clear |
|  | 1 = Specified |
| Type of location where study conducted and type of data collection performed | 0 = Not clear |
|  | 1 = Clearly stated |
| Sample size justification and power analysis | 0 = Unclear/not provided |
|  | 1 = Sufficiently described and justified before the study |

| **SCORE RANGE** | **RATING** |
| --- | --- |
| 0 to 3 | Low |
| 4 to 6 | Moderate |
| 7 to 9 | High |
